# Metabolic dysfunction-associated steatohepatitis is associated with increased all-cause mortality

**DOI:** 10.1101/2024.06.28.24309687

**Authors:** Zhao Li, Rui Song, Yingzhi Zhang, Jiahe Tan, Zhiwei Chen

## Abstract

**Background:** Recently, the new nomenclature metabolic dysfunction-associated steatohepatitis (MASH) was proposed to supersede non-alcoholic steatohepatitis (NASH). To optimize the management of these patients, it is crucial to comprehend the similarities and differences between individuals with NASH and MASH.

**Methods:** We analyzed data from 13,846 participants in the third National Health and Nutrition Examination Surveys, along with their linked mortality through 2019. NASH and MASH were defined based on respective criteria. Survey-weight adjusted multivariable Cox proportional model was used to examine mortality.

**Results:** The overall prevalence of steatohepatitis, NASH and MASH was 5.7% (n=788), 4.1% (n=564) and 5.5% (n=763), respectively. Most individuals with NASH (96.8%) could be categorized as MASH, but only 69.7% individuals with MASH qualified as NASH. During a median follow-up of 27 years, individuals with MASH exhibited a 53% higher risk of all-cause mortality (adjusted hazard ratio [aHR] 1.53, 95% CI 1.24-1.89). But individuals with NASH didn’t show an association with all-cause mortality after adjustment for metabolic risk factors (aHR 1.14, 95% CI 0.91-1.44). Notably, individuals who met the criteria for MASH but not NASH (NASH(-)/MASH(+)) had a higher risk of all-cause mortality (aHR 2.47, 95% CI 1.71-3.57) compared to those with NASH(+)/MASH(+) (aHR 1.22, 95% CI 0.97-1.55). Moreover, advanced fibrosis was only associated with an increased risk of all-cause mortality in individuals with MASH, not NASH.

**Conclusions:** MASH, rather than NASH, was associated with an elevated risk of all-cause mortality after adjusting for metabolic risk factors. Well-designed prospective studies are needed to assess and validate our findings.

## Introduction

The most severe form of nonalcoholic fatty liver disease (NAFLD), non-alcoholic steatohepatitis (NASH), significantly heightens the risk of developing cirrhosis and liver cancer^1^. Although the prevalence of NASH is relatively low compared to NAFLD, it is witnessing a global upsurge^2^. Recently, the nomenclature for NAFLD has been updated to metabolic dysfunction associated steatotic liver disease (MASLD)^3^, with NASH now designated as metabolic dysfunction-associated steatohepatitis (MASH). However, whether MASH exhibits similar characteristics or clinical outcomes as NASH remains ambiguous. Consequently, it is imperative to address these questions to optimize management strategies for individuals with MASH.

Compared with NAFLD, MASLD places greater emphasis on the influence of metabolic factors (meeting at least one of the five cardio-metabolic adult criteria, without the constraints of alcohol consumption or viral hepatitis)^3^. Our previous study, as well as those of others, have demonstrated a high concurrence between NAFLD and MASLD^4–8^. Nevertheless, the similarities and differences between NASH and MASH remain obscure. As per the definition^3^, it is speculated that individuals who met NASH definition but not MASH might lack or exhibit mild metabolic risk factors, whereas, those with MASH but not NASH are likely to present with more metabolic risk factors, heavier alcohol consumption, or coexistence of viral hepatitis (hepatitis B or hepatitis C). Although the incidence of discordant subgroups that fulfill the criteria for one but not the other between NASH and MASH might be rare^9^, concrete data is currently lacking.

A previous investigation utilizing the data from the Third National Health and Nutrition Examination Survey (NHANES III) reported that NASH was not associated with an elevated mortality rate over a 15-year follow-up period^10^. However, the potential impact of extending the follow-up duration on the outcomes remains undefined. Moreover, it is unclear whether MASH or the discordant subgroups exhibit differing risks of all-cause and cause-specific mortality compared to NASH, necessitating further exploration.

In this study, we employed data from NHANES III, with a 27-year follow-up period, to compare the similarities and differences in clinical features and long-term mortality between individuals with non-alcoholic steatohepatitis and metabolic dysfunction-associated steatohepatitis.

## Participants and Methods

### Study participants

In this study, we utilized data from the NHANES III (1988-1994), a comprehensive, stratified, clustered, multi-staged probability sampling design national survey that accurately represents the entire US population^11^. The outcomes of interviews, physical examinations, and laboratory tests on participants with available liver ultrasonography data were collected and analyzed. This mainly encompassed demographic indices (e.g., sex, age, race, education, smoking or drinking status, concomitant disease etc.), anthropometric indices (e.g., body mass index (BMI), waist circumference, etc.), and laboratory tests (e.g., alanine aminotransferase (ALT), aspartate aminotransferase (AST), and triglyceride, etc.). The NHANES mortality follow-up was a prospective study that dynamically assessed the vital status of all participants (≥20 years). Cause-specific and all-cause mortality data were obtained from the National Death Index up to December 2019^12^. Detailed information on data collection and mortality evaluation is provided in the supplementary materials. All individuals participating in NHANES III signed informed consent.

### Definition of NASH, MASH, and Fibrosis

As liver biopsies were not obtained in NHANES III, steatohepatitis was defined as mild to severe hepatic steatosis with elevated liver enzymes levels based on a previous study^10^. NASH was defined as the presence of steatohepatitis in the absence of excessive alcohol consumption (≥20 g/d for females and ≥30 g/d for males) and/or other liver diseases (such as viral hepatitis)^10^. MASH was defined by the presence of steatohepatitis in conjunction with at least one of the five cardio-metabolic adult criteria^3^. The detailed cardio-metabolic criteria are provided in the supplementary materials. We further categorized concordant NASH(+)/MASH(+) individuals as those who met both NASH and MASH definitions, while NASH(-)/MASH(-) individuals satisfied neither NASH nor MASH criteria. For the discordant subgroups, individuals fulfilling the NASH definition but not MASH were labeled as NASH(+)/MASH(-), and vice versa for those meeting MASH but not NASH, who were defined as NASH(-)/MASH(+). Finally, individuals without hepatic steatosis were designated as the control group.

Due to the unavailability of liver biopsy for direct assessment of fibrosis stage, we employed various non-invasive fibrosis scores as alternatives. The NAFLD fibrosis score (NFS) and fibrosis-4 (FIB-4) score were selected based on prior studies^13, 14^, which can be further categorized into low, intermediate, and high probabilities for advanced fibrosis. We defined advanced fibrosis as individuals presenting at least one of the two scores indicating a high probability of advanced fibrosis.

### Statistical analysis

The data are presented as median and interquartile range (IQR) for continuous variables, unweighted frequency counts and weighted percentage for categorical variables. The Wilcoxon test was employed for continuous variables, while the chi-square test was utilized for categorical variables. Cox proportional hazards regression was employed to estimate hazard ratios (HR) and 95% confidence intervals (CI) for deaths from all-causes and cause-specific (cardiovascular disease and cancer). We utilized two models with progressive degrees of adjustment: model 1 adjusted for demographic indices (sex, age, race, marital status, education, poverty impact ratio (PIR), healthy eating index (HEI) scores, sedentary lifestyle, and smoking status), while model 2 further adjusted for metabolic risk factors (BMI, waist circumference, triglycerides, and high-density lipoprotein cholesterol). For sensitivity analysis, firstly, to assess the impact of hepatic steatosis severity, we excluded individuals with mild hepatic steatosis from those with hepatic steatosis. Secondly, to avoid the potential confounding effect of viral hepatitis, we excluded participants with hepatitis B or hepatitis C from the entire population. All analyses were conducted using R (version 4.3.0; R Foundation for Statistical Computing, Vienna, Austria), with the survey package employed to account for the sampling weights and the complex survey design in the NHANES III.

## Results

### Characteristics of individuals with NASH or MASH

Of the 14,797 participants (20-74 years) who underwent hepatic/gallbladder ultrasound examinations in the NHANES III, 941 individuals were excluded due to ungradable or missing ultrasound images. In addition, 10 participants without follow-up data on mortality were also excluded. Consequently, 13,846 participants from NHANES III were included in this study (Table S1). The overall prevalence of steatohepatitis, NASH, and MASH were 5.7% (n=788), 4.1% (n=564), and 5.5% (n=763), respectively. To focus on steatohepatitis more effectively, individuals with hepatic steatosis and normal liver enzyme levels (n=4,225) were further excluded from the entire population. Ultimately, a total of 9,621 individuals were included for subsequent analyses. Compared to individuals without hepatic steatosis, those with steatohepatitis were more likely to be older, male, non-Hispanic black, less educated, poorer, have increased liver enzyme levels, and exhibit metabolic abnormalities (Table 1).

**Table 1.** Baseline characteristics of individuals with NASH or MASH.

| **Characteristics** | **No hepatic steatosis**<br>**(n=8,833)** | **Steatohepatitis**<br>**(n=788)** | **P** | **NASH**<br>**(n=564)** | **MASH**<br>**(n=763)** |
| --- | --- | --- | --- | --- | --- |
| Age (years) | 38.0 (28.0, 51.0) | 40.0 (33.0, 53.0) | 0.003 | 40.0 (32.0, 52.0) | 41.0 (33.0, 53.0) |
| Sex (male) | 4,027 (47%) | 445 (58%) | 0.007 | 290 (55%) | 429 (58%) |
| Race |  |  | <0.001 |  |  |
| Non-Hispanic white | 3,293 (76%) | 186 (66%) |  | 138 (70%) | 181 (67%) |
| Non-Hispanic black | 2,829 (12%) | 149 (8.6%) |  | 318 (14%) | 410 (14%) |
| Mexican-American | 2,324 (4.8%) | 421 (13%) |  | 89 (6.6%) | 141 (8.4%) |
| Others | 387 (7.7%) | 32 (12%) |  | 19 (9.8%) | 31 (11%) |
| Body mass index (kg/m<sup>2</sup>) | 24.7 (22.1, 27.6) | 30.0 (26.6, 35.1) | <0.001 | 30.6 (27.0, 35.6) | 30.3 (27.1, 35.2) |
| Waist circumference (cm) | 88 (79, 96) | 104 (94, 112) | <0.001 | 105 (94, 112) | 104 (94, 112) |
| ≥12 years education | 5,716 (79%) | 389 (70%) | <0.001 | 284 (74%) | 373 (69%) |
| Marital status | 5,445 (66%) | 493 (63%) | 0.3 | 374 (66%) | 484 (63%) |
| PIR | 2.9 (1.7, 4.3) | 2.3 (1.1, 3.8) | 0.016 | 2.4 (1.2, 3.9) | 2.2 (1.1, 3.9) |
| HEI score | 63 (54, 73) | 61 (53, 73) | 0.4 | 63 (53, 74) | 61 (53, 72) |
| Hypertension | 2,458 (23%) | 334 (49%) | <0.001 | 237 (47%) | 334 (51%) |
| Diabetes | 581 (3.9%) | 170 (20%) | <0.001 | 134 (21%) | 170 (21%) |
| Smoking status |  |  | 0.004 |  |  |
| Never | 4,341 (46%) | 377 (45%) |  | 310 (50%) | 363 (44%) |
| Past smoker | 1,824 (22%) | 195 (30%) |  | 148 (33%) | 191 (31%) |
| Current smoker | 2,668 (32%) | 216 (24%) |  | 106 (17%) | 209 (25%) |
| Alcohol consumption (g/d) | 4 (1, 12) | 8 (2, 28) | 0.003 | 3 (0, 10) | 8 (2, 28) |
| Viral hepatitis (HBV/HCV) | 242 (2.2%) | 96 (10%) | <0.001 | 0 | 96 (11%) |
| Sedentary lifestyle | 2,420 (19%) | 261 (23%) | 0.2 | 199 (24%) | 256 (23%) |
| Triglycerides (mg/dL) | 1.1 (0.8, 1.6) | 1.9 (1.2, 3.2) | <0.001 | 2.1 (1.4, 3.6) | 2.0 (1.3, 3.3) |
| HDL-cholesterol (mg/dL) | 1.3 (1.1, 1.6) | 1.1 (0.9, 1.4) | <0.001 | 1.0 (0.9, 1.3) | 1.1 (0.9, 1.4) |
| Glucose (mol/L) | 5.1 (4.8, 5.4) | 5.4 (5.1, 6.2) | <0.001 | 5.4 (5.0, 6.1) | 5.5 (5.1, 6.2) |
| ALT (U/L) | 13 (10, 18) | 47 (37, 63) | <0.001 | 47 (37, 63) | 47 (36, 63) |
| AST (U/L) | 18 (16, 22) | 39 (33, 53) | <0.001 | 38 (31, 48) | 39 (32, 53) |

Data are shown as the median (interquartile range) or unweighted frequency counts (weighted percentage) as appropriate. The Wilcoxon test for continuous variables and the Chi-square test for categorical variables were used in this analysis.
ALT, alanine aminotransferase; AST, aspartate aminotransferase; HDL, high-density lipoprotein; HEI, healthy eating index; MASH, metabolic dysfunction-associated steatohepatitis; NASH, nonalcoholic steatohepatitis; PIR, poverty impact ratio.

Of note, a significant majority individuals with NASH (96.8%) can be classified as MASH, yet only 69.7% of those with MASH can be qualified as NASH. Specifically, of the 788 individuals diagnosed with steatohepatitis, 69.3% of the subjects were assigned to the NASH(+)/MASH(+) subgroup, followed by NASH(-)/MASH(+), NASH(+)/MASH(-), and NASH(-)/MASH(-) subgroups (27.5%, 2.3%, and 0.9%, respectively; Figure 1). Notably, the subgroups characterized by MASH(+) exhibited elevated coexisting metabolic risk factors, while those characterized by NASH(-) demonstrated higher alcohol consumption and proportion of viral hepatitis, particularly in the NASH(-)/MASH(+) subgroup (Table S2).

**Figure 1.**
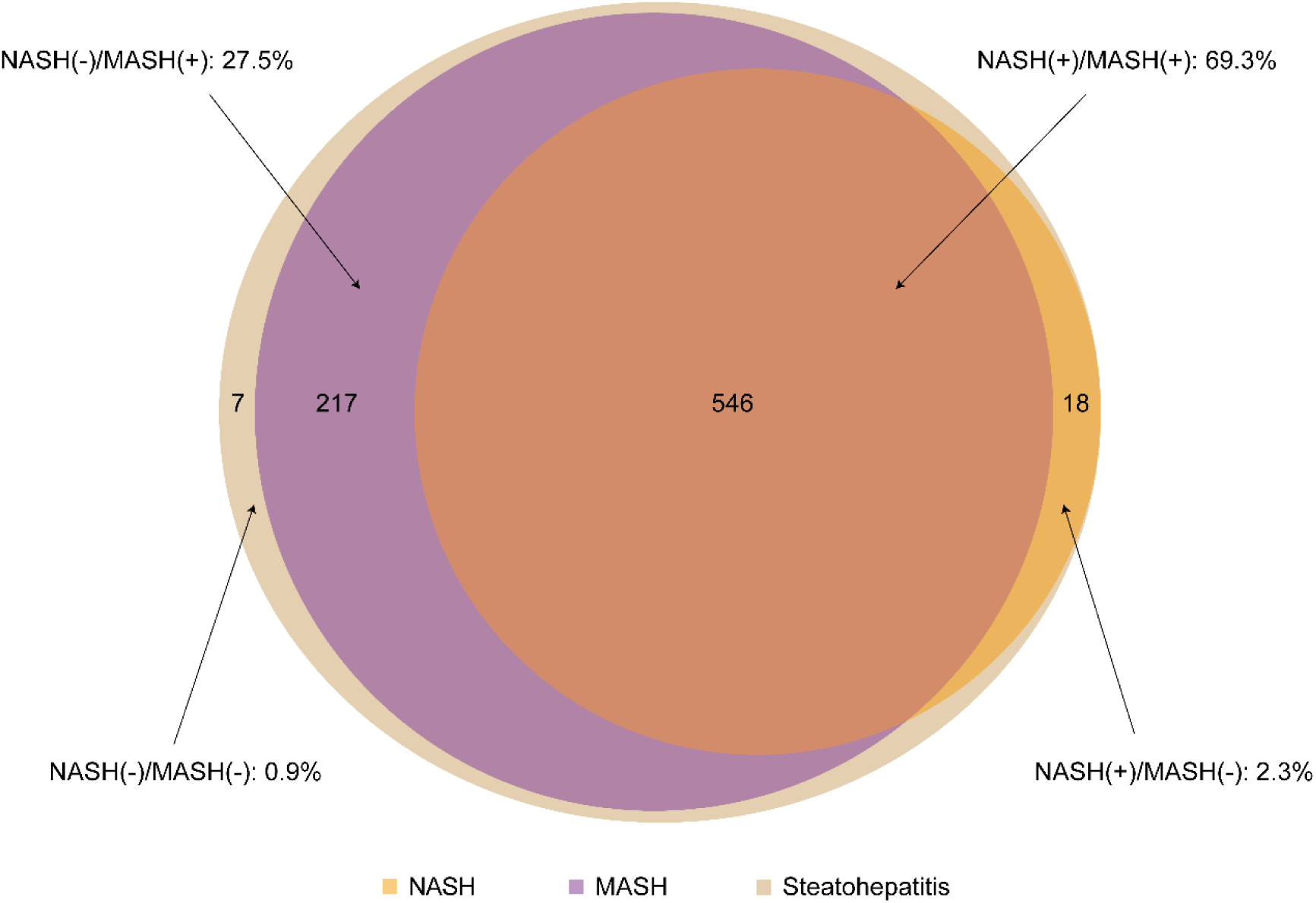
Relative proportions of non-alcoholic steatohepatitis and metabolic dysfunction-associated steatohepatitis. NASH(+)/MASH(+) was defined as individuals who met both NASH and MASH definitions. NASH(-)/MASH(-) was defined as individuals who met neither NASH nor MASH definitions. For the discordant subgroups, individuals meeting the definition of NASH but not MASH were defined as NASH(+)/MASH(-), while those meeting with MASH but not NASH were defined as NASH(-)/MASH(+).

### All-cause and cause-specific mortality of individuals with NASH or MASH

During the median follow-up period of 27.0 years (23.3-29.0), the cumulative mortality rate from all-cause was 32.3% (3,105 deaths). Cause-specific mortality was attributed to cardiovascular disease and cancer, accounting for 8.5% (816 deaths) and 8.1% (777 deaths), respectively. As shown in Table 2, individuals with NASH exhibited a 1.3-fold increased risk of all-cause mortality compared to those without NASH (HR 1.29, 95% CI 1.01-1.65). After adjusting for demographic and traditional risk factors, the association between NASH and all-cause mortality remained significant (Model 1: adjusted HR [aHR] 1.41, 95% CI 1.14-1.74). However, this association was attenuated when further adjusting for metabolic risk factors (Model 2: aHR 1.14, 95% CI 0.91-1.42). Notably, individuals with MASH exhibited higher all-cause mortality rate than those without MASH in both models (Model 1: aHR 1.74, 95% CI 1.44-2.1; Model 2: aHR 1.53, 95% CI 1.24-1.89). Intriguingly, subgroup analysis revealed that only the NASH(-)/MASH(+) and NASH(-)/MASH(-) subgroups demonstrated significantly elevated all-cause mortality rates compared to individuals without hepatic steatosis in model 2 (aHR 2.47, 95% CI 1.71-3.57; aHR 3.65, 95% CI 1.48-9.04). The concordant NASH(+)/MASH(+) subgroup showed no association with all-cause mortality after further adjusting for metabolic risk factors (aHR 1.22, 95% CI 0.97-1.55). (Table 2) Regarding cause-specific mortality, neither NASH nor MASH, nor their combined subgroups demonstrated an association with cardiovascular mortality (Model 2: aHR 0.74-1.43) or cancer mortality (Model 2: aHR 1.25-1.87; Table 3).

**Table 2.** Association among NASH or MASH status and all-cause mortality.

|  | N | n<br>(deaths) | Univariable model |  | Multivariable Model 1 |  | Multivariable Model 2 |  |
| --- | --- | --- | --- | --- | --- | --- | --- | --- |
|  |  |  | HR (95% CI) | P | HR (95% CI) | P | HR (95% CI) | P |
| No NASH | 9057 | 2900 | 1 |  | 1 |  | 1 |  |
| NASH | 564 | 205 | 1.29 (1.01-1.65) | 0.042 | 1.41 (1.14-1.74) | 0.001 | 1.14 (0.91-1.44) | 0.2 |
| No MASH | 8858 | 2792 | 1 |  | 1 |  | 1 |  |
| MASH | 763 | 313 | 1.8 (1.45-2.23) | <0.001 | 1.74 (1.44-2.1) | <0.001 | 1.53 (1.24-1.89) | <0.001 |
| No hepatic steatosis | 8833 | 2785 | 1 |  | 1 |  | 1 |  |
| NASH(+)/MASH(+) | 546 | 203 | 1.41 (1.08-1.82) | 0.01 | 1.47 (1.18-1.82) | <0.001 | 1.22 (0.97-1.55) | 0.1 |
| NASH(+)/MASH(-) | 18 | 2 | 0.11 (0.02-0.61) | 0.012 | 0.41 (0.07-2.44) | 0.3 | 0.29 (0.03-2.77) | 0.3 |
| NASH(-)/MASH(+) | 217 | 110 | 3.12 (2.39-4.06) | <0.001 | 2.51 (1.77-3.56) | <0.001 | 2.47 (1.71-3.57) | <0.001 |
| NASH(-)/MASH(-) | 7 | 5 | 4.02 (1.56-10.4) | 0.004 | 3.01 (1.27-7.16) | 0.012 | 3.65 (1.48-9.04) | 0.005 |

Survey-weight adjusted multivariable Cox proportional models were used in this analysis.
Multivariate model 1 was adjusted for sex, age, race, marital status, education, PIR, HEI scores, sedentary lifestyle, and smoking status.
Multivariate model 2 was further adjusted for BMI, waist circumference, triglycerides, and high-density lipoprotein cholesterol in addition to model 1.
BMI, body mass index; CI, confidential intervals; HR, hazard ratio; HEI, healthy eating index; MASH, metabolic dysfunction-associated steatohepatitis; NASH, nonalcoholic steatohepatitis; PIR, poverty impact ratio.

**Table 3.**
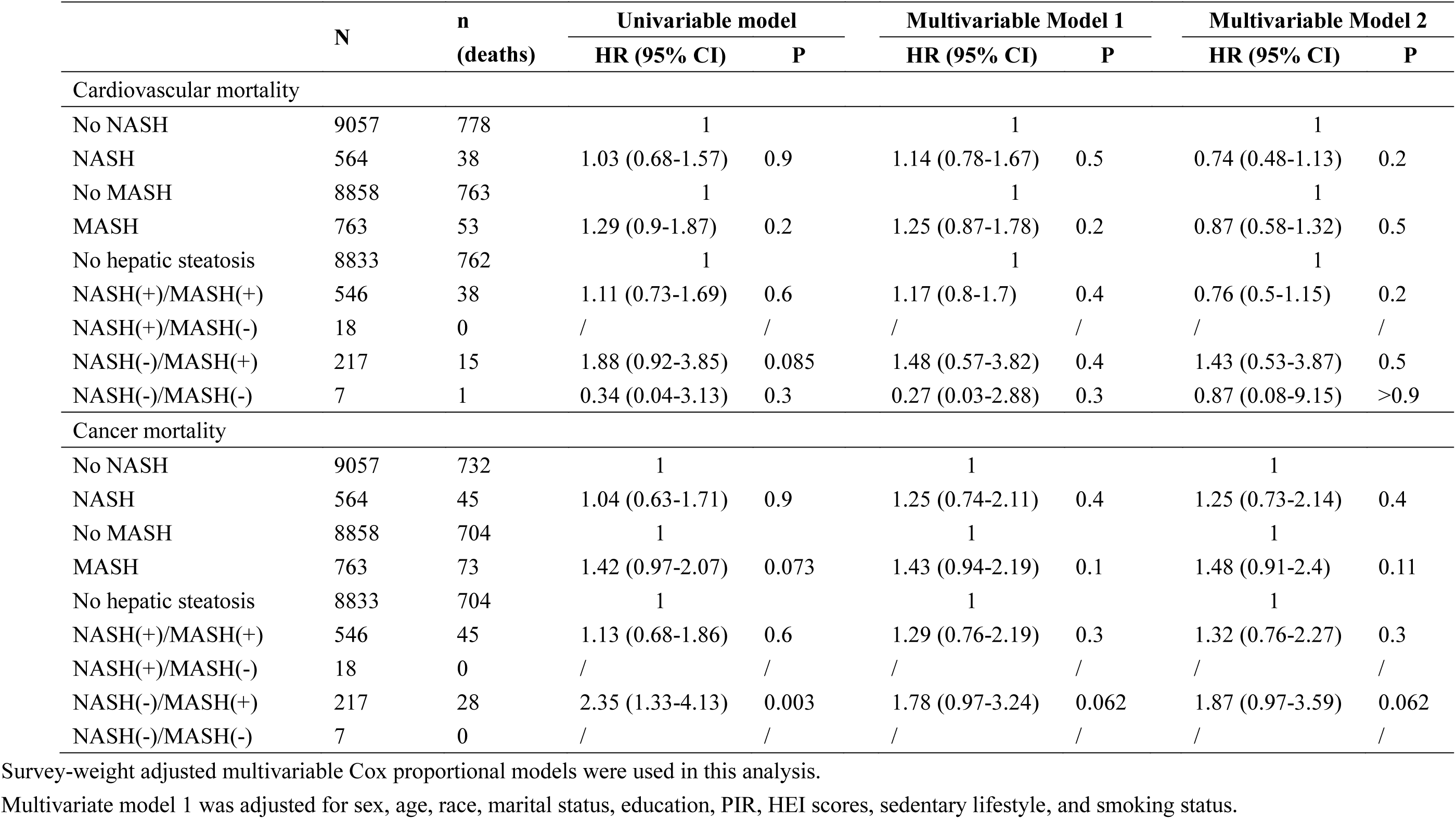
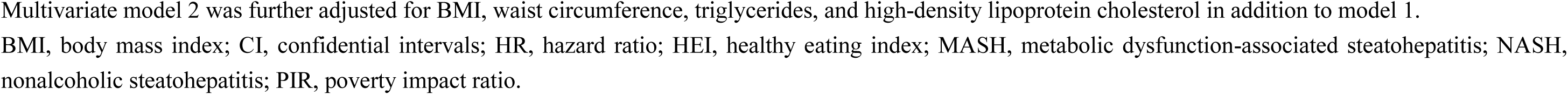
Association among NASH or MASH status and cardiovascular disease and cancer-related mortality.

|  | N | n<br>(deaths) | Univariable model |  | Multivariable Model 1 |  | Multivariable Model 2 |  |
| --- | --- | --- | --- | --- | --- | --- | --- | --- |
|  |  |  | HR (95% CI) | P | HR (95% CI) | P | HR (95% CI) | P |
| Cardiovascular mortality |  |  |  |  |  |  |  |  |
| No NASH | 9057 | 778 | 1 |  | 1 |  | 1 |  |
| NASH | 564 | 38 | 1.03 (0.68-1.57) | 0.9 | 1.14 (0.78-1.67) | 0.5 | 0.74 (0.48-1.13) | 0.2 |
| No MASH | 8858 | 763 | 1 |  | 1 |  | 1 |  |
| MASH | 763 | 53 | 1.29 (0.9-1.87) | 0.2 | 1.25 (0.87-1.78) | 0.2 | 0.87 (0.58-1.32) | 0.5 |
| No hepatic steatosis | 8833 | 762 | 1 |  | 1 |  | 1 |  |
| NASH(+)/MASH(+) | 546 | 38 | 1.11 (0.73-1.69) | 0.6 | 1.17 (0.8-1.7) | 0.4 | 0.76 (0.5-1.15) | 0.2 |
| NASH(+)/MASH(-) | 18 | 0 | / | / | / | / | / | / |
| NASH(-)/MASH(+) | 217 | 15 | 1.88 (0.92-3.85) | 0.085 | 1.48 (0.57-3.82) | 0.4 | 1.43 (0.53-3.87) | 0.5 |
| NASH(-)/MASH(-) | 7 | 1 | 0.34 (0.04-3.13) | 0.3 | 0.27 (0.03-2.88) | 0.3 | 0.87 (0.08-9.15) | >0.9 |
| Cancer mortality |  |  |  |  |  |  |  |  |
| No NASH | 9057 | 732 | 1 |  | 1 |  | 1 |  |
| NASH | 564 | 45 | 1.04 (0.63-1.71) | 0.9 | 1.25 (0.74-2.11) | 0.4 | 1.25 (0.73-2.14) | 0.4 |
| No MASH | 8858 | 704 | 1 |  | 1 |  | 1 |  |
| MASH | 763 | 73 | 1.42 (0.97-2.07) | 0.073 | 1.43 (0.94-2.19) | 0.1 | 1.48 (0.91-2.4) | 0.11 |
| No hepatic steatosis | 8833 | 704 | 1 |  | 1 |  | 1 |  |
| NASH(+)/MASH(+) | 546 | 45 | 1.13 (0.68-1.86) | 0.6 | 1.29 (0.76-2.19) | 0.3 | 1.32 (0.76-2.27) | 0.3 |
| NASH(+)/MASH(-) | 18 | 0 | / | / | / | / | / | / |
| NASH(-)/MASH(+) | 217 | 28 | 2.35 (1.33-4.13) | 0.003 | 1.78 (0.97-3.24) | 0.062 | 1.87 (0.97-3.59) | 0.062 |
| NASH(-)/MASH(-) | 7 | 0 | / | / | / | / | / | / |

Survey-weight adjusted multivariable Cox proportional models were used in this analysis.
Multivariate model 1 was adjusted for sex, age, race, marital status, education, PIR, HEI scores, sedentary lifestyle, and smoking status.
Multivariate model 2 was further adjusted for BMI, waist circumference, triglycerides, and high-density lipoprotein cholesterol in addition to model 1.
BMI, body mass index; CI, confidential intervals; HR, hazard ratio; HEI, healthy eating index; MASH, metabolic dysfunction-associated steatohepatitis; NASH, nonalcoholic steatohepatitis; PIR, poverty impact ratio.

### The effect of fibrosis on all-cause mortality in individuals with NASH or MASH

Given that individuals with NASH have a higher propensity to develop fibrosis than those with NAFLD^15^, we ponder whether such fibrosis augments the risk of all-cause mortality in NASH or MASH patients. Interestingly, advanced fibrosis was merely associated with an elevated risk of all-cause mortality in individuals with MASH, but not NASH (Model 2: aHR 1.7, 95% CI 1.12-2.58, p=0.012; aHR 1.39, 95% CI 0.86-2.22, p=0.2; Table 4). In particular, both the NFS and FIB-4 scores demonstrated a graded increase in the risk of all-cause mortality in individuals with MASH. That is, compared to the low probability subgroup, the high probability of advanced fibrosis subgroup exhibited a higher risk of mortality than the intermediate probability subgroup (NFS score: aHR 2.05, 95% CI 1.1-3.83; aHR 3.23, 95% CI 1.32-7.88; FIB-4 score: aHR 1.42, 95% CI 0.87-2.32; aHR 2.62, 95% CI 1.26-5.41). A similar trend was also observed in the NFS score of individuals with NASH (aHR 2.12, 95% CI 1.06-4.25; aHR 2.32, 95% CI 1.02-5.3), whereas, no such discrepancy was detected in the FIB-4 score. (Table 4)

**Table 4.** Association of advanced fibrosis status and all-cause mortality among individuals with NASH or MASH.

|  | N | n<br>(deaths) | Univariable model |  | Multivariable Model 1 |  | Multivariable Model 2 |  |
| --- | --- | --- | --- | --- | --- | --- | --- | --- |
|  |  |  | HR (95% CI) | P | HR (95% CI) | P | HR (95% CI) | P |
| NASH |  |  |  |  |  |  |  |  |
| No advanced fibrosis | 503 | 161 | 1 |  | 1 |  | 1 |  |
| Advanced fibrosis | 55 | 43 | 4.39 (2.68-7.19) | <0.001 | 1.51 (0.92-2.47) | 0.1 | 1.39 (0.86-2.22) | 0.2 |
| Low NFS | 377 | 78 | 1 |  | 1 |  | 1 |  |
| Intermediate NFS | 141 | 93 | 4.85 (2.60-9.03) | <0.001 | 2.65 (1.41-4.99) | 0.002 | 2.12 (1.06-4.25) | 0.033 |
| High NFS | 40 | 33 | 7.69 (3.55-16.7) | <0.001 | 3.11 (1.62-5.97) | <0.001 | 2.32 (1.02-5.30) | 0.046 |
| Low FIB-4 | 394 | 92 | 1 |  | 1 |  | 1 |  |
| Intermediate FIB-4 | 123 | 79 | 3.88 (2.31-6.52) | <0.001 | 1.45 (0.81-2.61) | 0.2 | 1.35 (0.66-2.73) | 0.4 |
| High FIB-4 | 41 | 33 | 8.87 (5.36-14.7) | <0.001 | 1.62 (0.75-3.50) | 0.2 | 1.95 (0.94-4.06) | 0.074 |
| MASH |  |  |  |  |  |  |  |  |
| No advanced fibrosis | 665 | 236 | 1 |  | 1 |  | 1 |  |
| Advanced fibrosis | 91 | 75 | 4.37 (3.18-6.01) | <0.001 | 1.92 (1.27-2.91) | 0.002 | 1.70 (1.12-2.58) | 0.012 |
| Low NFS | 499 | 123 | 1 |  | 1 |  | 1 |  |
| Intermediate NFS | 199 | 139 | 3.84 (2.38-6.19) | <0.001 | 2.55 (1.46-4.45) | <0.001 | 2.05 (1.10-3.83) | 0.024 |
| High NFS | 58 | 49 | 7.38 (4.21-12.9) | <0.001 | 4.40 (2.21-8.73) | <0.001 | 3.23 (1.32-7.88) | 0.01 |
| Low FIB-4 | 501 | 126 | 1 |  | 1 |  | 1 |  |
| Intermediate FIB-4 | 180 | 121 | 3.05 (2.12-4.39) | <0.001 | 1.38 (0.89-2.14) | 0.15 | 1.42 (0.87-2.32) | 0.2 |
| High FIB-4 | 75 | 64 | 8.01 (5.29-12.1) | <0.001 | 2.32 (1.26-4.27) | 0.007 | 2.62 (1.26-5.41) | 0.01 |

Survey-weight adjusted multivariable Cox proportional models were used in this analysis.
Multivariate model 1 was adjusted for sex, age, race, marital status, education, PIR, HEI scores, sedentary lifestyle, and smoking status.
Multivariate model 2 was further adjusted for BMI, waist circumference, triglycerides, and high-density lipoprotein cholesterol in addition to model 1.
BMI, body mass index; CI, confidential intervals; FIB-4, fibrosis-4; HR, hazard ratio; HEI, healthy eating index; MASH, metabolic dysfunction-associated steatohepatitis; NASH, nonalcoholic steatohepatitis; NFS, NAFLD fibrosis score; PIR, poverty impact ratio.

### Sensitivity analyses for individuals with NASH or MASH

First, we wonder whether the hepatic steatosis degrees could affect the clinical outcomes. After excluded individuals with mild hepatic steatosis from those with hepatic steatosis, similar findings of mortality outcomes were observed (Table S3-5). Second, to avoid the potential confounding effect of viral hepatitis, we excluded individuals with hepatitis B or hepatitis C from the whole population. In this scenario, the increased all-cause mortality was also observed in individuals with MASH, and NASH(-)/MASH(+) and NASH(-)/MASH(-) subgroups (Table S6). For the cause-specific mortality, the cardiovascular and cancer mortality were also not associated with both NASH or MASH and their combinations (Table S7). Even though the high NFS and FIB-4 scores subgroups still showed higher risk of all-cause mortality among individuals with MASH, the difference was not significant in increased all-cause mortality in MASH individuals with advanced fibrosis, after excluded participants with viral hepatitis (Model 2: aHR 1.41, 95% CI 0.99-2.03, p=0.059; Table S8).

Firstly, we examined the potential impact of hepatic steatosis severity on clinical outcomes. By excluding individuals with mild hepatic steatosis from those with hepatic steatosis, we observed similar mortality outcomes (Table S3-5). Secondly, to mitigate the potential confounding influence of viral hepatitis, we excluded individuals with hepatitis B or hepatitis C from the study population. In this context, increased all-cause mortality was also noted in individuals with MASH, as well as in the NASH(-)/MASH(+) and NASH(-)/MASH(-) subgroups (Table S6). Regarding cause-specific mortality, cardiovascular and cancer mortality were not associated with both NASH and MASH, nor with their combinations (Table S7). Although individuals with MASH in the high NFS and FIB-4 score subgroups exhibit a higher risk of all-cause mortality, the increased risk of all-cause mortality among MASH patients with advanced fibrosis does not reach statistical significance, after excluding participants with viral hepatitis (Model 2: aHR 1.41, 95% CI 0.99-2.03, p=0.059; Table S8).

## Discussion

The recent proposal of the new nomenclature MASH to replace NASH necessitates a thorough understanding of the similarities and differences between individuals with NASH and MASH, in order to optimize the management of these patients. In this study, although patients with NASH exhibited similar clinical features regarding metabolic risk factors compared to those with MASH, it was found that MASH, not NASH, was significantly associated with an increased all-cause mortality after adjusting for metabolic risk factors. Additionally, advanced fibrosis in patients with MASH was associated with a higher risk of all-cause mortality compared to those with NASH. These findings imply that independent of metabolic risk factors, there exists a strong correlation between MASH and an increase in all-cause mortality.

NASH can contribute to progressive fibrosis, and in some cases, cirrhosis or even liver cancer^1^. However, a previous study found no correlation between NASH and an increase in all-cause mortality following a 15-year follow-up in NHANES III^10^. Using a longer follow-up period (27 years), we reiterated that NASH per se is not associated with all-cause mortality after adjusting for metabolic risk factors. Notably, in contrast to the previous study^10^, NASH increases the risk of all-cause mortality when only adjusting for demographic and traditional risk factors (model 1), but not metabolic risk factors. Although the definition of NASH does not encompass metabolic risk, individuals with NASH exhibit similar metabolic characteristics to those with MASH in this study. Consequently, caution should be exercised for the long-term prognosis of individuals with NASH, particularly those with a high risk of concurrent conditions (e.g., type 2 diabetes mellitus^16^ or liver cirrhosis^17^). Nevertheless, individuals with MASH exhibit a 53% higher risk of all-cause mortality than those without MASH even after adjusting for metabolic risks. This suggests that MASH can identify individuals with a higher risk of all-cause mortality than NASH. Therefore, greater attention and early intervention are warranted for those with MASH, such as lifestyle modification^18, 19^, or other novel therapies^20, 21^.

When we further categorized individuals with steatohepatitis into four subgroups, interesting findings regarding all-cause mortality emerged. In Model 1, individuals with NASH(+)/MASH(+) exhibited a 47% increased risk of all-cause mortality, which disappeared after further adjusting for metabolic risk factors. This suggests that metabolic factors play a significant role in the elevation of all-cause mortality risk for these individuals. Notably, individuals with NASH(-)/MASH(+) still exhibited a 2.47-fold risk of all-cause mortality even after adjusting for metabolic risk factors. This indicates that, beyond metabolic factors, heavy alcohol consumption and/or viral hepatitis may have a more potent impact on the risk of all-cause mortality for MASH patients^22, 23^. After excluding individuals with viral hepatitis in the sensitivity analysis, the high all-cause mortality risk persisted. This suggests that significant alcohol consumption, rather than viral hepatitis, could be responsible for the increased all-cause mortality risk. However, this should be interpreted with caution due to the small sample size. Larger studies are needed to verify these findings in the future. A similar phenomenon was also observed in individuals with NASH(-)/MASH(-), although the sample size was smaller. This subgroup represents a relatively small proportion of individuals with concurrent liver disease but without metabolic comorbidities. After excluding individuals with viral hepatitis, this discordant group may be attributed to alcoholic steatohepatitis (ASH), which could contribute to the interpretation of the increased risk of all-cause mortality^24^. Therefore, based on these observations, it is crucial to distinguish between MASH with concurrent liver disease and its absence in future studies.

In this study, a significant majority of patients with NASH (96.8%) are classified as MASH, with only 3.2% lacking metabolic risk factors. This finding is consistent with a previous review^9^, and supports the retention and validity of historical data from NASH clinical trials. However, the situation becomes distinct when reversed. As it is not a prerequisite to exclude concurrent liver diseases for the diagnosis of MASH, MASH may encompass a broader range of individuals with various liver disease etiologies (viral, alcoholic, autoimmune, genetic, etc.), akin to MASLD^3^. Specifically, only 69.7% of individuals with MASH can be confirmed as having NASH, suggesting that 30.3% of these individuals harbor alternative etiologies (excessive alcohol consumption and/or viral hepatitis) in this analysis. The gap may vary with differing population selection. Moreover, in this context, it appears challenging to differentiate between MASH and hepatitis induced by other etiologies (e.g., ASH or active viral hepatitis). This could potentially impact the risk of all-cause mortality for these patients directly, as mentioned above. From this aspect, it may be advantageous to appropriately refine or qualify the new term of MASH in order to mitigate this ambiguity.

To our knowledge, no study has previously evaluated the association between MASH and mortality. This is the strength of our study. However, there are several limitations to our study. First, although this is a nationally representative study based on the NHANES III dataset (1988-1994), the low prevalence of steatohepatitis at that time resulted in a relatively small sample of patients with NASH or MASH. Large-sample studies focusing on these patients are needed to validate our observations. Second, steatohepatitis was not confirmed by liver biopsy but by ultrasound examination, which may underestimate the prevalence and impact on clinical outcomes of individuals with NASH or MASH^25^. However, the advantages of ultrasound, such as safety, affordability, sufficient sensitivity, and satisfactory specificity, make it suitable for use in large population-based epidemiologic studies^26^. Third, in the US population-based study, the prevalence of viral hepatitis (especially hepatitis B) was low, therefore, the results in this regard should be interpreted with caution. Further studies are needed to clarify the specific effect of viral hepatitis on the mortality of individuals with MASH, especially in high-prevalence areas of viral hepatitis, such as Asia and Africa^27^.

In summary, our research demonstrates for the first time a significant association between MASH and increased all-cause mortality, independently of metabolic and demographic risk factors. In contrast, NASH exhibits a heightened risk of all-cause mortality, while it becomes non-significant following adjustment for metabolic risk factors. Moreover, it is crucial to differentiate between individuals with concomitant liver disease and those without when studying MASH. In future, well-designed prospective studies with large sample sizes are needed to assess and validate our findings.

## Acknowledgement

We gratefully acknowledge the contribution of the participants of the NHANES cohort, research assistants, and facilitating personnel. And this work was supported by the Science and technology research project of Chongqing Education Commission (KJZD-K202300404), Remarkable Innovation-Clinical Research Project and the DengFeng Program of the Second Affiliated Hospital of Chongqing Medical University, and The First batch of key Disciplines on Public Health in Chongqing, Health Commission of Chongqing, China.

## Ethics approval statement

The National Center for Health Statistics Research Ethics Review Board approved the NHANES protocol.

## Data Availability Statement

All data produced in the present study are available upon reasonable request to the authors.

## Author contributions

Rui Song, Zhao Li and Yingzhi Zhang was involved in acquisition of data, data analysis and interpretation, drafting of the manuscript, and study supervision. Jiahe Tan were involved in interpretation of data and critical revision of the manuscript. Zhiwei Chen was involved in study concept and design, interpretation of data, drafting of the manuscript, critical revision of the manuscript, and study supervision. All authors contributed to the article and approved the submitted version.

## Data Availability Statement

All the data in this study were available in the NHANES III public database.

## Ethics approval statement and patient consent statement

NHANES III was approved by the CDC/NCHS Ethics Review Board, and all individuals signed informed consent to participate in NHANES III.

## Permission to reproduce material from other sources

Yes.

## Clinical trial registration

Not applicable.

## Abbreviations

ALT: alanine aminotransferase
AST: aspartate aminotransferase
BMI: body mass index
CI: confidential intervals
FIB-4: fibrosis-4
HR: hazard ratio
HEI: healthy eating index
MASH: metabolic dysfunction-associated steatohepatitis
MASLD: metabolic dysfunction associated steatotic liver disease
NAFLD: nonalcoholic fatty liver disease
NASH: nonalcoholic steatohepatitis
NHANES III: Third National Health and Nutrition Examination Survey
NFS: NAFLD fibrosis score
PIR: poverty impact ratio.

