## Supplementary material for "Metabolic dysfunction-associated steatohepatitis is associated with increased all-cause mortality": Tables: Supplementary materials.docx

**Supplementary methods**

**Data collection**

The demographic characteristics (e.g., sex, age, ethnicity, education, marital status, smoking, alcohol consumption, physical activity, underlying diseases, and drug use), anthropometric indices (e.g., body mass index (BMI), waist circumference, and blood pressure), and laboratory test results (e.g., alanine aminotransferase, aspartate aminotransferase, HDL-cholesterol, and fasting glucose) of the participants were retrieved. The definitions of various factors, such as hypertension, diabetes, smoking status, alcohol consumption, and sedentary lifestyle, were provided as follows.

Hypertension was defined as systolic blood pressure ≥140 mm Hg or diastolic blood pressure ≥90 mm Hg, and/or self-reported physician diagnosis, and/or use of antihypertensive medication^1^. Diabetes mellitus was defined as fasting glucose ≥126 mg/dl, hemoglobin A1c ≥6.5%, and/or self-reported physician diagnosis, and/or treatment with antidiabetic medication^2^. Smoking status was categorized as never, past, and current smokers. Never smokers were those who answered no to the question: “Have you smoked at least 100 cigarettes during your entire life?” Current smokers were defined as individuals who reported ongoing smoking and had smoked at least 100 cigarettes in their lifetime^2^. Alcohol consumption was calculated as “grams of alcohol intake per day (g/d)”. In detail, the average daily number of alcoholic drinks was estimated by multiplying the number of drinking days over the past 12 months and the average number of drinks on a drinking day, and dividing by 365^1^. According to US standards, alcoholic drinks were counted as 14 g of alcohol each ^3^. In addition, never drinkers were those who answered no to the question: “In your entire life, have you had at least 12 drinks of any kind of alcoholic beverage?” Average daily alcohol consumption was defined as zero for never drinkers in this study. Excessive alcohol consumption was defined as ≥20 g/d for females and ≥30 g/d for males^4^. A sedentary lifestyle was defined as individuals who answered ’no’ to all questions about engaging in any of the following physical activities over the last month: jogging or running, cycling, swimming, aerobics, other dancing, calisthenics, garden or yard work, weight lifting, or other sports^1^. Hepatitis B virus (HBV) was defined as hepatitis B surface antigen-positive. Hepatitis C virus (HCV) was defined as hepatitis C antibody-positive.

**All-cause and cause-specific mortality**

In NHANES III, mortality was assessed utilizing the Underlying Cause of Death 113 (UCOD_113) code. All-cause mortality was classified according to the ICD-9 for deaths through 1998, and transitioned to ICD-10 for deaths from 1999 to 2019. Cause-specific mortalities were examined, focusing on cardiovascular disease (UCOD_113 code: 55-64, 70) and cancer (UCOD_113 code: 19-43)^2^. Furthermore, we calculated the follow-up time (months) from the date of interview to the date of death or the end of the follow-up period.

**Definition of SLD, NAFLD, and MASLD**

We defined steatosis liver diseases (SLD) as mild to severe hepatic steatosis without increased liver enzymes levels in this study. NAFLD was defined as the presence of SLD without excessive alcohol consumption (≥20 g/d for females and ≥30 g/d for males) and/or other liver diseases (such as viral hepatitis) ^5^. MASLD was defined as the presence of SLD combined with at least 1 of the 5 following cardiometabolic adult criteria: (1) BMI ≥25 kg/m^2^ or waist circumference ≥94 cm for males and ≥80 cm for females, (2) fasting glucose ≥100 mg/dl or 2-hour post-load glucose levels ≥140mg/dl or hemoglobin A1c ≥5.7% or diabetes mellitus or treatment for diabetes mellitus, (3) blood pressure ≥130/85 mmHg or antihypertensive drug treatment, (4) fasting plasma triglycerides ≥150 mg/dl or lipid-lowering treatment, (5) plasma HDL-cholesterol <40 mg/dl for men and <50 mg/dl for women or lipid-lowering treatment ^4^.

**Table S1. Baseline characteristics of the entire population (N=13,846).**

| **Characteristics** | **All individuals** |
| --- | --- |
| Age (years) | 40.0 (30.0, 53.0) |
| Sex (male) | 6,475 (48%) |
| Race |  |
| Non-Hispanic white | 5,032 (75%) |
| Non-Hispanic black | 4,157 (5.5%) |
| Mexican-American | 4,076 (11%) |
| Others | 387 (7.7%) |
| Body mass index (kg/m^2^) | 25.5 (22.6, 29.4) |
| Waist circumference (cm) | 91 (81, 101) |
| ≥12 years education | 8,446 (77%) |
| Marital status | 8,769 (67%) |
| PIR | 2.8 (1.6, 4.3) |
| HEI score | 63 (54, 73) |
| Hypertension | 4,543 (28%) |
| Diabetes | 1,514 (7.1%) |
| Smoking status |  |
| Never | 6,726 (45%) |
| Past smoker | 3,179 (25%) |
| Current smoker | 3,940 (30%) |
| Alcohol consumption (g/d) | 4 (1, 12) |
| Viral hepatitis (HBV/HCV) | 408 (2.5%) |
| Sedentary lifestyle | 4,139 (21%) |
| Triglycerides (mg/dL) | 1.2 (0.9, 1.9) |
| HDL-cholesterol (mg/dL) | 1.2 (1.0, 1.5) |
| Glucose (mol/L) | 5.1 (4.8, 5.5) |
| ALT (U/L) | 14 (10, 21) |
| AST (U/L) | 19 (16, 23) |

Data are shown as the median (interquartile range) or unweighted frequency counts (weighted percentage) as appropriate.

ALT, alanine aminotransferase; AST, aspartate aminotransferase; HDL, high-density lipoprotein; HEI, healthy eating index; MASH, metabolic dysfunction-associated steatohepatitis; NASH, nonalcoholic steatohepatitis; PIR, poverty impact ratio.

**Table S2. Baseline characteristics of the final study population (N = 9,621).**

| **Characteristics** | **No hepatic steatosis**  **(n=8,833)** | **NASH(+)**  **MASH(+)**  **(n=546)** | **NASH(+)**  **MASH(-)**  **(n=18)** | **NASH(-)**  **MASH(+)**  **(n=217)** | **NASH(-)**  **MASH(-)**  **(n=7)** | **P** |
| --- | --- | --- | --- | --- | --- | --- |
| Age (years) | 38.0 (28.0, 51.0) | 40.0 (33.0, 52.0) | 32.0 (27.0, 36.1) | 43.0 (36.0, 56.0) | 51.1 (31.9, 54.7) | 0.001 |
| Sex (male) | 4,027 (47%) | 279 (55%) | 11 (62%) | 150 (66%) | 5 (39%) | <0.001 |
| Race/ethnicity |  |  |  |  |  | <0.001 |
| Non-Hispanic white | 3,293 (76%) | 134 (70%) | 4 (55%) | 47 (57%) | 1 (49%) |  |
| Non-Hispanic black | 2,829 (12%) | 82 (6.4%) | 7 (12%) | 59 (14%) | 1 (20%) |  |
| Mexican-American | 2,324 (4.8%) | 312 (14%) | 6 (5.2%) | 98 (12%) | 5 (31%) |  |
| Others | 387 (7.7%) | 18 (8.8%) | 1 (28%) | 13 (17%) | 0 (0%) |  |
| Body mass index (kg/m^2^) | 24.7 (22.1, 27.6) | 30.8 (27.6, 36.0) | 22.9 (19.6, 24.1) | 28.5 (26.0, 33.5) | 19.9 (19.4, 21.6) | <0.001 |
| Waist circumference (cm) | 88 (79, 96) | 105 (95, 112) | 79 (71, 85) | 101 (91, 111) | 77 (76, 77) | <0.001 |
| ≥12 years education | 5,716 (79%) | 272 (73%) | 12 (93%) | 101 (59%) | 4 (87%) | <0.001 |
| Marital status | 5,445 (66%) | 368 (67%) | 6 (52%) | 116 (54%) | 3 (23%) | 0.11 |
| PIR | 2.9 (1.7, 4.3) | 2.4 (1.2, 3.9) | 2.7 (2.4, 2.9) | 1.8 (0.9, 3.4) | 0.5 (0.1, 1.8) | <0.001 |
| HEI score | 63 (54, 73) | 63 (53, 73) | 66 (48, 76) | 60 (52, 70) | 67 (56, 70) | 0.002 |
| Hypertension | 2,458 (23%) | 237 (49%) | 0 (0%) | 97 (54%) | 0 | <0.001 |
| Diabetes | 581 (3.9%) | 134 (22%) | 0 (0%) | 36 (19%) | 0 | <0.001 |
| Smoking status |  |  |  |  |  | <0.001 |
| Never | 4,341 (46%) | 298 (48%) | 12 (71%) | 65 (33%) | 2 (57%) |  |
| Past smoker | 1,824 (22%) | 145 (34%) | 3 (24%) | 46 (23%) | 1 (12%) |  |
| Current smoker | 2,668 (32%) | 103 (18%) | 3 (5.4%) | 106 (44%) | 4 (31%) |  |
| Alcohol consumption (g/d) | 4 (1, 12) | 3 (0, 11) | 3 (2, 3) | 40 (24, 60) | 27 (25, 34) | <0.001 |
| Viral hepatitis (HBV/HCV) | 242 (2.2%) | 0 | 0 | 96 (38%) | 0 | <0.001 |
| Sedentary lifestyle | 2,420 (19%) | 194 (24%) | 5 (25%) | 62 (20%) | 0 | <0.001 |
| Triglycerides (mg/dL) | 1.1 (0.8, 1.6) | 2.2 (1.6, 3.6) | 1.0 (0.6, 1.2) | 1.4 (0.9, 2.5) | 0.8 (0.7, 0.9) | <0.001 |
| HDL-cholesterol (mg/dL) | 1.3 (1.1, 1.6) | 1.0 (0.9, 1.2) | 1.5 (1.3, 1.6) | 1.4 (1.1, 1.6) | 3.0 (2.5, 3.6) | <0.001 |
| Glucose (mol/L) | 5.1 (4.8, 5.4) | 5.4 (5.0, 6.2) | 5.2 (4.9, 5.2) | 5.7 (5.2, 6.3) | 4.6 (4.5, 4.7) | <0.001 |
| ALT (U/L) | 13 (10, 18) | 48 (37, 63) | 45 (36, 78) | 46 (31, 64) | 71 (18, 78) | <0.001 |
| AST (U/L) | 18 (16, 22) | 38 (30, 48) | 39 (37, 47) | 45 (38, 63) | 104 (37, 115) | <0.001 |

Data are shown as the median (interquartile range) or unweighted frequency counts (weighted percentage) as appropriate. The Kruskal-Wallis test for continuous variables and the Chi-square test for categorical variables were used in this analysis.

ALT, alanine aminotransferase; AST, aspartate aminotransferase; HDL, high-density lipoprotein; HEI, healthy eating index; MASH, metabolic dysfunction-associated steatohepatitis; NASH, nonalcoholic steatohepatitis; PIR, poverty impact ratio.

**Table S3. Association among NASH or MASH status and all-cause mortality (for the sensitivity analysis 1^#^).**

|  | **N** | **n**  **(deaths)** | **Univariable model** | |  | **Multivariable Model 1** | |  | **Multivariable Model 2** | |
| --- | --- | --- | --- | --- | --- | --- | --- | --- | --- | --- |
|  |  |  | **HR (95% CI)** | **P** |  | **HR (95% CI)** | **P** |  | **HR (95% CI)** | **P** |
| No NASH | 8992 | 2873 | 1 | |  | 1 | |  | 1 | |
| NASH | 461 | 171 | 1.4 (1.06-1.84) | 0.017 |  | 1.44 (1.13-1.83) | 0.003 |  | 1.16 (0.89-1.51) | 0.3 |
| No MASH | 8850 | 2792 | 1 | |  | 1 | |  | 1 | |
| MASH | 603 | 252 | 1.79 (1.42-2.25) | <0.001 |  | 1.69 (1.36-2.11) | <0.001 |  | 1.45 (1.15-1.84) | 0.002 |
| No hepatic steatosis | 8833 | 2785 | 1 | |  | 1 | |  | 1 | |
| NASH(+)/MASH(+) | 450 | 169 | 1.47 (1.11-1.93) | 0.006 |  | 1.47 (1.16-1.87) | 0.002 |  | 1.22 (0.93-1.59) | 0.15 |
| NASH(+)/MASH(-) | 11 | 2 | 0.24 (0.04-1.3) | 0.1 |  | 1.27 (0.25-6.53) | 0.8 |  | 0.93 (0.11-8.15) | >0.9 |
| NASH(-)/MASH(+) | 153 | 83 | 3.06 (2.25-4.17) | <0.001 |  | 2.47 (1.65-3.7) | <0.001 |  | 2.34 (1.51-3.63) | <0.001 |
| NASH(-)/MASH(-) | 6 | 5 | 4.65 (1.83-11.8) | 0.001 |  | 3.06 (1.27-7.41) | 0.013 |  | 3.61 (1.45-9.02) | 0.006 |

^#^ (n=9,453) excluding individuals with mild hepatic steatosis.

Survey-weight adjusted multivariable Cox proportional models were used in this analysis.

Multivariate model 1 was adjusted for sex, age, race, marital status, education, PIR, HEI scores, sedentary lifestyle, and smoking status.

Multivariate model 2 was further adjusted for BMI, waist circumference, triglycerides, and high-density lipoprotein cholesterol in addition to model 1.

BMI, body mass index; CI, confidential intervals; HR, hazard ratio; HEI, healthy eating index; MASH, metabolic dysfunction-associated steatohepatitis; NASH, nonalcoholic steatohepatitis; PIR, poverty impact ratio.

**Table S4. Association among NASH or MASH status and cardiovascular disease and cancer-related mortality (for the sensitivity analysis 1^#^).**

|  | **N** | **n**  **(deaths)** | **Univariable model** | |  | **Multivariable Model 1** | |  | **Multivariable Model 2** | |
| --- | --- | --- | --- | --- | --- | --- | --- | --- | --- | --- |
|  |  |  | **HR (95% CI)** | **P** |  | **HR (95% CI)** | **P** |  | **HR (95% CI)** | **P** |
| Cardiovascular mortality |  |  |  | |  |  | |  |  | |
| No NASH | 8992 | 776 | 1 | |  | 1 | |  | 1 | |
| NASH | 461 | 34 | 1.17 (0.75-1.83) | 0.5 |  | 1.26 (0.85-1.86) | 0.3 |  | 0.78 (0.50-1.21) | 0.3 |
| No MASH | 8850 | 763 | 1 | |  | 1 | |  | 1 | |
| MASH | 603 | 47 | 1.49 (1.04-2.12) | 0.029 |  | 1.41 (0.98-2.05) | 0.067 |  | 0.95 (0.62-1.47) | 0.8 |
| No hepatic steatosis | 8833 | 762 | 1 | |  | 1 | |  | 1 | |
| NASH(+)/MASH(+) | 450 | 34 | 1.23 (0.8-1.89) | 0.3 |  | 1.28 (0.87-1.88) | 0.2 |  | 0.81 (0.53-1.23) | 0.3 |
| NASH(+)/MASH(-) | 11 | 0 | / | / |  | / | / |  | / | / |
| NASH(-)/MASH(+) | 153 | 13 | 2.5 (1.12-5.58) | 0.025 |  | 1.92 (0.65-5.67) | 0.2 |  | 1.8 (0.58-5.59) | 0.3 |
| NASH(-)/MASH(-) | 6 | 1 | 0.39 (0.04-3.85) | 0.4 |  | 0.27 (0.03-2.86) | 0.3 |  | 0.89 (0.08-9.39) | >0.9 |
| Cancer mortality |  |  |  |  |  |  |  |  |  |  |
| No NASH | 8992 | 724 | 1 |  |  | 1 |  |  | 1 |  |
| NASH | 461 | 37 | 0.99 (0.57-1.74) | >0.9 |  | 1.12 (0.63-1.99) | 0.7 |  | 1.09 (0.61-1.95) | 0.8 |
| No MASH | 8850 | 704 | 1 |  |  | 1 |  |  | 1 |  |
| MASH | 603 | 57 | 1.22 (0.76-1.96) | 0.4 |  | 1.27 (0.8-2.03) | 0.3 |  | 1.27 (0.76-2.11) | 0.4 |
| No hepatic steatosis | 8833 | 704 | 1 |  |  | 1 |  |  | 1 |  |
| NASH(+)/MASH(+) | 450 | 37 | 1.03 (0.59-1.81) | >0.9 |  | 1.14 (0.64-2.03) | 0.7 |  | 1.12 (0.62-2.02) | 0.7 |
| NASH(+)/MASH(-) | 11 | 0 | / | / |  | / | / |  | / | / |
| NASH(-)/MASH(+) | 153 | 20 | 1.96 (0.91-4.19) | 0.084 |  | 1.7 (0.84-3.41) | 0.14 |  | 1.72 (0.81-3.65) | 0.2 |
| NASH(-)/MASH(-) | 6 | 0 | / | / |  | / | / |  | / | / |

^#^ (n=9,453) excluding individuals with mild hepatic steatosis.

Survey-weight adjusted multivariable Cox proportional models were used in this analysis.

Multivariate model 1 was adjusted for sex, age, race, marital status, education, PIR, HEI scores, sedentary lifestyle, and smoking status.

Multivariate model 2 was further adjusted for BMI, waist circumference, triglycerides, and high-density lipoprotein cholesterol in addition to model 1.

BMI, body mass index; CI, confidential intervals; HR, hazard ratio; HEI, healthy eating index; MASH, metabolic dysfunction-associated steatohepatitis; NASH, nonalcoholic steatohepatitis; PIR, poverty impact ratio.

**Table S5. Association of advanced fibrosis status and all-cause mortality among individuals with NASH or MASH (for the sensitivity analysis 1^#^).**

|  | **N** | **n (deaths)** | **Univariable model** | |  | **Multivariable Model 1** | |  | **Multivariable Model 2** | |
| --- | --- | --- | --- | --- | --- | --- | --- | --- | --- | --- |
|  |  |  | **HR (95% CI)** | **P** |  | **HR (95% CI)** | **P** |  | **HR (95% CI)** | **P** |
| **NASH** |  |  |  |  |  |  |  |  |  |  |
| No advanced fibrosis | 412 | 137 | 1 | |  | 1 | |  | 1 | |
| Advanced fibrosis | 43 | 33 | 3.93 (2.10-7.38) | <0.001 |  | 1.68 (0.94-3.00) | 0.079 |  | 1.35 (0.79-2.30) | 0.3 |
| Low NFS | 303 | 67 | 1 | |  | 1 | |  | 1 | |
| Intermediate NFS | 120 | 78 | 3.51 (1.78-6.94) | <0.001 |  | 2.15 (1.13-4.09) | 0.019 |  | 1.48 (0.78-2.81) | 0.2 |
| High NFS | 32 | 25 | 5.82 (2.42-14.0) | <0.001 |  | 2.73 (1.43-5.24) | 0.002 |  | 1.62 (0.76-3.46) | 0.2 |
| Low FIB-4 | 320 | 77 | 1 | |  | 1 | |  | 1 | |
| Intermediate FIB-4 | 105 | 69 | 3.81 (1.99-7.30) | <0.001 |  | 1.64 (0.82-3.29) | 0.2 |  | 1.51 (0.65-3.52) | 0.3 |
| High FIB-4 | 30 | 24 | 7.88 (4.45-14.0) | <0.001 |  | 1.93 (0.76-4.90) | 0.2 |  | 2.19 (0.88-5.42) | 0.09 |
| **MASH** |  |  |  |  |  |  |  |  |  |  |
| No advanced fibrosis | 523 | 191 | 1 | |  | 1 | |  | 1 | |
| Advanced fibrosis | 73 | 59 | 4.19 (2.75-6.39) | <0.001 |  | 1.93 (1.20-3.10) | 0.007 |  | 1.55 (1.01-2.38) | 0.043 |
| Low NFS | 390 | 102 | 1 | |  | 1 | |  | 1 | |
| Intermediate NFS | 159 | 110 | 3.39 (2.04-5.64) | <0.001 |  | 2.42 (1.34-4.37) | 0.003 |  | 1.71 (0.89-3.28) | 0.11 |
| High NFS | 47 | 38 | 6.34 (3.22-12.5) | <0.001 |  | 3.88 (1.89-7.97) | <0.001 |  | 2.35 (0.96-5.75) | 0.062 |
| Low FIB-4 | 395 | 105 | 1 | |  | 1 | |  | 1 | |
| Intermediate FIB-4 | 143 | 96 | 2.98 (1.82-4.87) | <0.001 |  | 1.45 (0.84-2.50) | 0.2 |  | 1.4 (0.77-2.55) | 0.3 |
| High FIB-4 | 58 | 49 | 7.52 (4.72-12.0) | <0.001 |  | 2.43 (1.16-5.09) | 0.019 |  | 2.54 (1.14-5.65) | 0.022 |

^#^ (n=9,453) excluding individuals with mild hepatic steatosis.

Survey-weight adjusted multivariable Cox proportional models were used in this analysis.

Multivariate model 1 was adjusted for sex, age, race, marital status, education, PIR, HEI scores, sedentary lifestyle, and smoking status.

Multivariate model 2 was further adjusted for BMI, waist circumference, triglycerides, and high-density lipoprotein cholesterol in addition to model 1.

BMI, body mass index; CI, confidential intervals; FIB-4, fibrosis-4; HR, hazard ratio; HEI, healthy eating index; MASH, metabolic dysfunction-associated steatohepatitis; NASH, nonalcoholic steatohepatitis; NFS, NAFLD fibrosis score; PIR, poverty impact ratio.

**Table S6. Association among NASH or MASH status and all-cause mortality (for the sensitivity analysis 2^#^).**

|  | **N** | **n**  **(deaths)** | **Univariable model** | |  | **Multivariable Model 1** | |  | **Multivariable Model 2** | |
| --- | --- | --- | --- | --- | --- | --- | --- | --- | --- | --- |
|  |  |  | **HR (95% CI)** | **P** |  | **HR (95% CI)** | **P** |  | **HR (95% CI)** | **P** |
| No NASH | 8719 | 2735 | 1 | |  | 1 | |  | 1 | |
| NASH | 564 | 205 | 1.32 (1.03-1.7) | 0.029 |  | 1.45 (1.17-1.8) | <0.001 |  | 1.2 (0.95-1.52) | 0.13 |
| No MASH | 8616 | 2678 | 1 | |  | 1 | |  | 1 | |
| MASH | 667 | 262 | 1.67 (1.32-2.11) | <0.001 |  | 1.62 (1.3-2.03) | <0.001 |  | 1.41 (1.13-1.76) | 0.003 |
| No hepatic steatosis | 8591 | 2671 | 1 | |  | 1 | |  | 1 | |
| NASH(+)/MASH(+) | 546 | 203 | 1.42 (1.1-1.85) | 0.008 |  | 1.49 (1.2-1.86) | <0.001 |  | 1.25 (0.99-1.59) | 0.065 |
| NASH(+)/MASH(-) | 18 | 2 | 0.11 (0.02-0.62) | 0.013 |  | 0.43 (0.07-2.58) | 0.4 |  | 0.31 (0.03-2.91) | 0.3 |
| NASH(-)/MASH(+) | 121 | 59 | 2.94 (2.1-4.13) | <0.001 |  | 2.15 (1.33-3.47) | 0.002 |  | 2.08 (1.28-3.37) | 0.003 |
| NASH(-)/MASH(-) | 7 | 5 | 4.08 (1.57-10.6) | 0.004 |  | 3.15 (1.33-7.46) | 0.009 |  | 3.85 (1.54-9.63) | 0.004 |

^#^ (n=9,283) excluding individuals with viral hepatitis.

Survey-weight adjusted multivariable Cox proportional models were used in this analysis.

Multivariate model 1 was adjusted for sex, age, race, marital status, education, PIR, HEI scores, sedentary lifestyle, and smoking status.

Multivariate model 2 was further adjusted for BMI, waist circumference, triglycerides, and high-density lipoprotein cholesterol in addition to model 1.

BMI, body mass index; CI, confidential intervals; HR, hazard ratio; HEI, healthy eating index; MASH, metabolic dysfunction-associated steatohepatitis; NASH, nonalcoholic steatohepatitis; PIR, poverty impact ratio.

**Table S7. Association among NASH or MASH status and cardiovascular disease and cancer-related mortality (for the sensitivity analysis 2^#^).**

|  | **N** | **n**  **(deaths)** | **Univariable model** | |  | **Multivariable Model 1** | |  | **Multivariable Model 2** | |
| --- | --- | --- | --- | --- | --- | --- | --- | --- | --- | --- |
|  |  |  | **HR (95% CI)** | **P** |  | **HR (95% CI)** | **P** |  | **HR (95% CI)** | **P** |
| Cardiovascular mortality |  |  |  | |  |  | |  |  | |
| No NASH | 8719 | 752 | 1 | |  | 1 | |  | 1 | |
| NASH | 564 | 38 | 1.05 (0.69-1.6) | 0.8 |  | 1.18 (0.81-1.72) | 0.4 |  | 0.74 (0.48-1.16) | 0.2 |
| No MASH | 8616 | 744 | 1 | |  | 1 | |  | 1 | |
| MASH | 667 | 46 | 1.21 (0.84-1.73) | 0.3 |  | 1.24 (0.89-1.72) | 0.2 |  | 0.82 (0.55-1.22) | 0.3 |
| No hepatic steatosis | 8591 | 743 | 1 | |  | 1 | |  | 1 | |
| NASH(+)/MASH(+) | 546 | 38 | 1.13 (0.75-1.7) | 0.6 |  | 1.2 (0.83-1.74) | 0.3 |  | 0.76 (0.49-1.16) | 0.2 |
| NASH(+)/MASH(-) | 18 | 0 | / | / |  | / | / |  | / | / |
| NASH(-)/MASH(+) | 121 | 8 | 1.6 (0.62-4.17) | 0.3 |  | 1.4 (0.42-4.65) | 0.6 |  | 1.32 (0.36-4.92) | 0.7 |
| NASH(-)/MASH(-) | 7 | 1 | 0.34 (0.04-3.15) | 0.3 |  | 0.28 (0.03-2.98) | 0.3 |  | 1.1 (0.11-11.1) | >0.9 |
| Cancer mortality |  |  |  |  |  |  |  |  |  |  |
| No NASH | 8719 | 697 | 1 |  |  | 1 |  |  | 1 |  |
| NASH | 564 | 45 | 1.04 (0.64-1.71) | 0.9 |  | 1.24 (0.74-2.09) | 0.4 |  | 1.25 (0.74-2.13) | 0.4 |
| No MASH | 8616 | 681 | 1 |  |  | 1 |  |  | 1 |  |
| MASH | 667 | 61 | 1.44 (0.96-2.15) | 0.074 |  | 1.44 (0.92-2.28) | 0.11 |  | 1.5 (0.9-2.51) | 0.12 |
| No hepatic steatosis | 8591 | 681 | 1 |  |  | 1 |  |  | 1 |  |
| NASH(+)/MASH(+) | 546 | 45 | 1.13 (0.68-1.86) | 0.6 |  | 1.28 (0.76-2.17) | 0.4 |  | 1.32 (0.77-2.27) | 0.3 |
| NASH(+)/MASH(-) | 18 | 0 | / | / |  | / | / |  | / | / |
| NASH(-)/MASH(+) | 121 | 16 | 3.01 (1.61-5.63) | <0.001 |  | 2 (1-3.99) | 0.049 |  | 2.11 (1-4.48) | 0.051 |
| NASH(-)/MASH(-) | 7 | 0 | / | / |  | / | / |  | / | / |

^#^ (n=9,283) excluding individuals with viral hepatitis.

Survey-weight adjusted multivariable Cox proportional models were used in this analysis.

Multivariate model 1 was adjusted for sex, age, race, marital status, education, PIR, HEI scores, sedentary lifestyle, and smoking status.

Multivariate model 2 was further adjusted for BMI, waist circumference, triglycerides, and high-density lipoprotein cholesterol in addition to model 1.

BMI, body mass index; CI, confidential intervals; HR, hazard ratio; HEI, healthy eating index; MASH, metabolic dysfunction-associated steatohepatitis; NASH, nonalcoholic steatohepatitis; PIR, poverty impact ratio.

**Table S8. Association of advanced fibrosis status and all-cause mortality among individuals with NASH or MASH (for the sensitivity analysis 2^#^).**

|  | **N** | **n (deaths)** | **Univariable model** | |  | **Multivariable Model 1** | |  | **Multivariable Model 2** | |
| --- | --- | --- | --- | --- | --- | --- | --- | --- | --- | --- |
|  |  |  | **HR (95% CI)** | **P** |  | **HR (95% CI)** | **P** |  | **HR (95% CI)** | **P** |
| **NASH** |  |  |  |  |  |  |  |  |  |  |
| No advanced fibrosis | 503 | 161 | 1 | |  | 1 | |  | 1 | |
| Advanced fibrosis | 55 | 43 | 4.39 (2.68-7.19) | <0.001 |  | 1.51 (0.92-2.47) | 0.1 |  | 1.39 (0.86-2.22) | 0.2 |
| Low NFS | 377 | 78 | 1 | |  | 1 | |  | 1 | |
| Intermediate NFS | 141 | 93 | 4.85 (2.60-9.03) | <0.001 |  | 2.65 (1.41-4.99) | 0.002 |  | 2.12 (1.06-4.25) | 0.033 |
| High NFS | 40 | 33 | 7.69 (3.55-16.7) | <0.001 |  | 3.11 (1.62-5.97) | <0.001 |  | 2.32 (1.02-5.30) | 0.046 |
| Low FIB-4 | 394 | 92 | 1 | |  | 1 | |  | 1 | |
| Intermediate FIB-4 | 123 | 79 | 3.88 (2.31-6.52) | <0.001 |  | 1.45 (0.81-2.61) | 0.2 |  | 1.35 (0.66-2.73) | 0.4 |
| High FIB-4 | 41 | 33 | 8.87 (5.36-14.7) | <0.001 |  | 1.62 (0.75-3.5) | 0.2 |  | 1.95 (0.94-4.06) | 0.074 |
| **MASH** |  |  |  |  |  |  |  |  |  |  |
| No advanced fibrosis | 589 | 201 | 1 | |  | 1 | |  | 1 | |
| Advanced fibrosis | 71 | 59 | 4.28 (3.05-6.00) | <0.001 |  | 1.69 (1.15-2.47) | 0.007 |  | 1.41 (0.99-2.03) | 0.059 |
| Low NFS | 437 | 99 | 1 | |  | 1 | |  | 1 | |
| Intermediate NFS | 176 | 122 | 3.84 (2.38-6.19) | <0.001 |  | 2.55 (1.46-4.45) | <0.001 |  | 2.05 (1.10-3.83) | 0.024 |
| High NFS | 47 | 39 | 7.38 (4.21-12.9) | <0.001 |  | 4.4 (2.21-8.73) | <0.001 |  | 3.23 (1.32-7.88) | 0.01 |
| Low FIB-4 | 449 | 108 | 1 | |  | 1 | |  | 1 | |
| Intermediate FIB-4 | 155 | 103 | 3.05 (2.06-4.52) | <0.001 |  | 1.24 (0.79-1.96) | 0.3 |  | 1.17 (0.72-1.89) | 0.5 |
| High FIB-4 | 56 | 49 | 8.39 (5.46-12.9) | <0.001 |  | 1.92 (1.02-3.6) | 0.042 |  | 1.91 (1.04-3.51) | 0.037 |

^#^ (n=9,283) excluding individuals with viral hepatitis.

Survey-weight adjusted multivariable Cox proportional models were used in this analysis.

Multivariate model 1 was adjusted for sex, age, race, marital status, education, PIR, HEI scores, sedentary lifestyle, and smoking status.

Multivariate model 2 was further adjusted for BMI, waist circumference, triglycerides, and high-density lipoprotein cholesterol in addition to model 1.

BMI, body mass index; CI, confidential intervals; FIB-4, fibrosis-4; HR, hazard ratio; HEI, healthy eating index; MASH, metabolic dysfunction-associated steatohepatitis; NASH, nonalcoholic steatohepatitis; NFS, NAFLD fibrosis score; PIR, poverty impact ratio.
